# Assessment of knowledge about human papillomavirus vaccination among primary school girls in Arba Minch town, South Ethiopia, 2020. An institution-based cross-sectional study

**DOI:** 10.1101/2021.10.16.21264889

**Authors:** Eshetu Yisihak Ukumo, Feleke Gebremeskel, Samuel Abebe, Desta Markos Minanmo, Gebresilasea Gendisha Ukke

## Abstract

**Introduction:** Cervical cancer is the second leading cause of cancer-related mortality among females in Ethiopia. The knowledge regarding human papillomavirus vaccination and its acceptability among adolescent girls affects the human papillomavirus vaccine uptake, however, the status of knowledge of the human papillomavirus vaccination among adolescent girls in Ethiopia, particularly in this study area is not well known. Therefore, this study aimed to determine the knowledge of human papillomavirus vaccination and associated factors among primary school girls in Arba Minch town, South Ethiopia, 2020.

**Methods:** A school-based cross-sectional study in which 516 school girls participated was conducted on January 24, 2020. The study participants were selected by a simple random sampling technique. A pre-tested and self-administered questionnaire was used to collect the data. Data was entered to Epi_data version 4.6 and exported to SPSS Version 23 for analysis. The logistic regression model was used to identify the statistically significant variables for knowledge of the human papillomavirus vaccination.

**Results:** The overall knowledge level of the study participants in this study was 71.7%. Their main information source was social media 41.74% followed by health care workers 29.69%.

**Conclusions:** Nearly three-fourths of the study participants were knowledgeable about human papillomavirus vaccination. The knowledge about the human papillomavirus vaccination shows a positive association with age, education level, and access to information sources, and parents’ educational status.

## Introduction

The human papillomavirus (HPV) vaccine is a vaccine used to prevent genital warts, anal cancer, cervical cancer, vulvar cancer, and vaginal cancer caused by certain types of human papillomavirus (HPV) [1]. Cervical cancer is a global public health problem accounting for almost 300,000 deaths annually. Eighty-three of new cases and 85% of related deaths occur in resource-poor countries [2]. Cervical cancer is the most notable health problem of Sub-Saharan Africa (SSA). Annually 34.8/100,000 women new cases and 22.5 / 100,000 women deaths were occurred in this sub-region [2]. In Ethiopia cervical cancer is the second leading cause of cancer mortality, every year, 7095 women are diagnosed and 4732 are dying from cervical cancer [3-5]. Of the totally served patients at the center, women were about 73 %, and cervical cancer is the most common disease comprising over one-third of all female patients treated in Ethiopia [6].

The majority of cancers (over 80%) in sub-Saharan Africa (SSA) are detected at a late stage, predominantly due to lack of information about cervical cancer and a scarcity of prevention services which in turn are associated with low survival rates after surgery or radiotherapy [7]. The primary prevention for cervical cancer is to avoid the transmission of HPV types. Among those methods, HPV vaccination is the one [7]. Similarly, with WHO, Ethiopia recommends primary target population; girls aged 9–14 years, for HPV vaccination before becoming sexually active.

Factors that are associated with the knowledge of HPV vaccine include information source [8], residence [9] family education level [10], age [11], school type, discussion with health care providers [10], School-based education about HPV vaccine [12], and Discussion about the vaccine with peer [13].

Even if knowledge regarding HPV vaccine among adolescent girls affects HPV vaccine uptake, knowledge of the HPV vaccine was little known in Ethiopia. Therefore, this study aimed to fill this information gap by assessing knowledge, and associated factors of HPV vaccine among primary school girls in Arbaminch town, south Ethiopia. The primary beneficiaries of this study were adolescent girls because their levels of knowledge towards the HPV vaccine was assessed and recommended to the concerned bodies. Secondly, the result of this study can work as a crucial source of information for health program planners and further investigators.

## MATERIALS AND METHODS

### Study setting

Arba-Minch town, the capital city of Gamo zone, is 505km from Addis Ababa (capital city of Ethiopia) and about 280 Km from Hawasa (capital city of southern Ethiopia). There are 1 hospital, 3 health centers, and 11 health posts providing health services. Arbaminch has 4 sub-cities and 11 kebeles. Arba Minch town has 55 KG schools from which 6 were public, 18 full primary schools from which 8 were public, 9 secondary and preparatory schools from which 5 were public. A total number of students registered in 2020 in Arbaminch town were grade one up to twelve 27,512 from which female students are 14,431, grade five up to eight 8,867 from which female students are 4,758, [14], zonal and city health offices work jointly with schools to deliver the vaccine in a school-based way after creating awareness by health extension workers and other concerned bodies [15].

### Study design

A school-based cross-sectional study

### Source population

All girl students in primary schools of Arba Minch town, South Ethiopia

### Study population

All girl students in 6 selected primary schools of Arba Minch town during the data collection period.

### Study variables

#### Dependent variable

Knowledge of HPV vaccine

#### Independent variables

Age,

Grade level,

School type,

Educational status of family,

Source of information about HPV vaccine

School based education about HPV vaccine

### Operational and term Definitions

**Good knowledge**, Those who answered 50% or more or 4 and above from 7 knowledge assessing questions [11].

**Poor knowledge**, Those who answered below (50%) from knowledge assessing questions [11].

**The human papillomavirus**; is a group of viruses that is extremely common worldwide [16].

**The human papillomavirus vaccine**; is a vaccine used to prevent cervical cancer, genital warts, anal cancers, caused by certain types of human papillomavirus [1]

### Eligibility criteria

#### Inclusion criteria

Those girls who were attending a primary school and in grade 5 up to 8

#### Exclusion criteria

Those girls who were ill at the time of data collection

#### Sample Size Determination

The sample size was determined by using a single population proportion formula with the assumption of a 95% confidence level, 5% margin of error, by taking “p” (0.696) from similar studies conducted among adolescent girls, design effect 1.5 and adding 10% non-response rate. The final sample size was 537.

#### Sampling technique

From total primary schools found in the city (in both private and public), 6 primary schools were selected by simple random sampling without replacement through the lottery method. Then the calculated sample size was allocated to each school proportionally, then to each grade level again proportionally, after getting eligible adolescent girls for each grade level from their homeroom teachers and relisted to prepare sampling frame to that grade level. while preparing the sampling frame, a section of given students was listed in front of their name to finally get her, then study units of a given grade level obtained by SRS through lottery method. Finally, selected study units for given schools were requested to be in one place (to one of the sections at break time) and fill the questionnaire through the guidance of data collection facilitators and supervisors.

#### Data collection tool and procedures

Selected girls were assembled in one place in (one of the sections at break time) selected schools on the day of the survey. A pre-tested, Semi-structured, translated to Amharic language and self-administered questionnaire was used. Each girl selected for the survey filled the questionnaire under close supervision of data collectors and investigators. Data collectors were selected from teachers of their respective schools. The questionnaire consists of socio-demographic information and knowledge of the HPV vaccine among adolescent girls of Arbaminch town. The tool was prepared by English version and translated to Amharic to avoid missing important information; then it was translated back to English for analysis.

#### Data quality control

The training was given on clarification of some assessment tools, the aim of the study, time of data collection, timely collection and reorganization of the collected data from respective schools, and submission on due time.

The questionnaire was pre-tested with 5% of the sample size in Mirab Abaya primary school in the Gamo zone which is not included in this study. In addition to appropriate recruitment and training of data collectors, the quality of the data was monitored frequently both in the field and during data entry. This was done in the field through close supervision of data collectors. All filled questionnaire was examined for completeness and consistency during data collection.

#### Data Processing and analysis

Data was entered to Epi data version 4.6 and exported for analysis to SPSS Version 23. Descriptive analysis was made and measures of central tendency were also determined. In scoring knowledge, one point was awarded for every correct answer and zero for incorrect or ‘do not know’ responses. The total knowledge score was then be converted into percentages and the level of knowledge of the respondents was classified based on their score and of overall 7 knowledge questions, study participants who score <50% (i.e. answered correctly ≤3 questions) were labeled as having poor knowledge and those who answer 4 or above from 7 questions or score 50% and above were labeled as having good knowledge.

Logistic regression was applied to see the association between dependent and independent variables. Independent variables, which had an association in bi-variable analysis with p-value <0.25 were entered into the multivariable logistic regression model. Independent variables with p-value<0.05 in the multivariable logistic regression model were considered as statically significant factors for knowledge. The results were presented as odds ratios (OR) with 95% confidence intervals. An odds ratio with corresponding 95% CI was used to quantify the association between a dependent variable and independent variables. The model fitness was checked by using the software application of the Hossmare and Lame show test (>0.05). The results were presented using text, and tables.

## Results

Out of the 537 samples that were distributed, 516 study participants were interviewed in this study, with a response rate of 96.1%. Majorities of the study participants were Gamo in ethnicity (71.3%) and protestant Christians in religion (60.5%) (Table 1).

**Table 1:** Socio-demographic characteristics of primary school girls in Arba Minch town, southern Ethiopia, 2020, (N=516).

| Variables | categories | Frequency (n) | Percent (%) |
| --- | --- | --- | --- |
| Age in years | 12_14 | 284 | 55 |
|  | ≥15 | 232 | 45 |
| Religion | Protestant | 312 | 60.5 |
|  | Orthodox | 148 | 28.7 |
|  | Catholic | 20 | 3.9 |
|  | Muslim | 17 | 3.3 |
|  | Others | 19 | 3.7 |
| Ethnic group | Gamo | 368 | 71.3 |
|  | Gofa | 46 | 8.9 |
|  | Amhara | 33 | 6.4 |
|  | Oromo | 23 | 4.5 |
|  | Wolayta | 33 | 6.4 |
|  | Others | 13 | 2.5 |
| Residence | Urban | 436 | 84.5 |
|  | Rural | 80 | 15.5 |
| Grade level | Grade 5&6 | 219 | 42.4 |
|  | Grade 7 &8 | 297 | 57.6 |
| School type | Public | 470 | 91.1 |
|  | Private | 46 | 8.9 |
| Mothers educational status | no education | 112 | 21.7 |
|  | primary education | 115 | 22.3 |
|  | secondary education | 184 | 35.7 |
|  | more than secondary | 105 | 20.3 |
| Fathers educational status | no education | 72 | 14.0 |
|  | primary education | 89 | 17.2 |
|  | secondary education | 107 | 20.7 |
|  | more than secondary | 248 | 48.1 |

### Knowledge of respondents toward HPV vaccine

Three hundred seventy, 71.7% CI 95%(67.6_75.4) of the study participants were found to have good knowledge about the HPV vaccine, 69.8% of the study participants knew the targeted age group and 78.3 knew the number of HPV vaccines to be given for prevention of cervical cancer. The main source of information was social media (41.74%) followed by health care providers while giving health information at school (29.69%) (Table2).

**Table 2:** Knowledge and related responses of primary school girls in Arba Minch town, south Ethiopia, 2020, (N=516).

| Variables |  | Frequency | Percent |
| --- | --- | --- | --- |
| Do you know HPV vaccine | Yes | 409 | 79.3 |
|  | No | 107 | 20.7 |
| What is HPV vaccine | Given for prevention of cervical cancer | 310 | 60.1 |
|  | Given for prevention of malaria | 50 | 9.7 |
|  | Given for prevention of HIV ADDIS | 16 | 3.1 |
|  | I do not know | 140 | 27.1 |
| An interval that someone take second HPV vaccine | 6 month | 314 | 60.9 |
|  | 6 year | 62 | 12.0 |
|  | 1 month | 26 | 5.0 |
|  | 3 month | 114 | 22.1 |
| HPV vaccines should be taken before sex initiations | Yes | 292 | 56.6 |
|  | No | 224 | 43.4 |
| Number of HPV vaccine | Twice | 404 | 78.3 |
|  | Once | 112 | 21.7 |
| Possible to have cervical cancer screening after HPV vaccination | Yes | 383 | 74.2 |
|  | No | 133 | 25.8 |
| Age at which someone undergo HPV vaccination | 9-14 years | 360 | 69.8 |
|  | 18-21 years | 30 | 5.8 |
|  | All ages | 27 | 5.2 |
|  | I do not know | 99 | 19.2 |
| Obtained information about HPV vaccine from any source | Yes | 357 | 69.2 |
|  | No | 159 | 30.8 |
| Information source | Social media | 149 | 41.74 |
|  | Health care workers | 106 | 29.69 |
|  | Families and friends | 102 | 28.57 |
| In your school, is there any reproductive health related clubs | Yes | 357 | 69.2 |
|  | No | 159 | 30.8 |
| Knowledge level | Poor Knowledge | 146 | 28.3 CI 95%(24.6_32.4) |
|  | Good Knowledge | 370 | 71.7 CI 95%(67.6_75.4) |

### Associated factors with knowledge of HPV vaccine

Logistic regression analysis was done to assess an association between the socio-demographic characteristics of the study participants with their knowledge level towards HPV vaccine showed that; age, Grade level, familial educational status, and information source, have had a positive association. (that is those who are >15 years old have close to fourfold more likely having good knowledge about HPV vaccine as compared to those who are 12_14 years old, AOR =3.74, 95%CI (2.20_6.37), P-value <0.001),

Those who are in Grade7and 8 close to four-fold more likely of having good knowledge about HPV vaccine as compared to those who are in Grade 5 and 6, AOR=3.98, 95%CI (2.40_6.58), P-value <0.001),

Familial educational status (those for whom, mothers educational status of secondary close to fourteen fold and more than secondary close to twenty twofold and fathers educational status of more than secondary close to threefold have good knowledge about HPV vaccine than their counterpart, AOR=13.60, 95%CI (5.69_32.53) P-value <0.001), 22.27, 95%CI(8.23_60.30), P-value<0.001), 2.18, 95%CI(1.09_4.35), P-value 0.03) respectively,

Those who obtained HPV vaccine information have close to nine fold more likely of having good knowledge about HPV vaccine than those who do not obtain HPV vaccine information AOR=8.65, 95% CI(3.92_19.07), P-value <0.001)) (table3). Hosmer and Lemeshow’s test value of this model was 0.131.

**Table 3:** Association between socio-demographic factors and knowledge level of HPV vaccine *= shows an association, ** = shows strong association

| Variables | Knowledge of HPV vaccine | COR(95% CI) | AOR(95% CI) |
| --- | --- | --- | --- |

|  |  | Good | Poor |  |  | P_value |
| --- | --- | --- | --- | --- | --- | --- |
| Age | 12_14 | 180(34.9%) | 104(20.2%) | 1 | 1 | 1 |
|  | ≥15 | 190(36.8%) | 42(8.2%) | 2.61(1.73_3.95) | 3.74(2.20_6.37) | <0.001** |
| School type | Public | 330(64%) | 140(27.1%) | 1 | 1 | 1 |
|  | Private | 40(7.8%) | 6(1.2%) | 2.83(1.17_6.82) | 1.704(0.6_4.87) | 0.32 |
| Residence | Rural | 52(10.1%) | 28(5.4%) | 1 | 1 | 1 |
|  | Urban | 318(61.6%) | 118(22.9%) | 1.45(0.88_2.41) | 1.27(0.68_2.39) | 0.46 |
| Grade level | 5 and 6 | 136(26.4%) | 83(16.1%) | 1 | 1 | 1 |
|  | 7and 8 | 234(45.3%) | 63(12.2%) | 2.27(1.54_3.35) | 3.98(2.40_6.58) | <0.001** |
| Mothers' educational status | No education | 64(12.4%) | 48(9.3%) | 1 | 1 | 1 |
|  | Primary | 58(11.2%) | 57(11%) | 0.76(0.45_1.29) | 0.94(0.50_1.76) | 0.84 |
|  | Secondary | 155(30%) | 29(5.6%) | 4.01(2.32_6.92) | 13.60(5.69_32.53) | <0.001** |
|  | More than secondary | 93(18%) | 12(2.3%) | 5.81(2.86_11.80) | 22.27(8.23_60.30) | <0.001** |
| Fathers' educational status | No education | 37(7.2%) | 35(6.8%) | 1 | 1 | 1 |
|  | Primary | 46(8.9%) | 43(8.3%) | 1.01(0.54_1.88) | 0.73(0.35_1.55) | 0.41 |
|  | Secondary | 83(16.1%) | 24(4.7%) | 3.27(1.71_6.25) | 1.98(0.90_4.36) | 0.09 |
|  | More than secondary | 204(39.5%) | 44(8.5%) | 4.39(2.49_7.72) | 2.18(1.09_4.35) | 0.03* |
| Herd about HPV vaccine from any source | Yes | 260(50.4%) | 97(18.8%) | 1 | 1 | 1 |
|  | No | 110(21.3%) | 49(9.5%) | 1.19(0.79_1.80) | 8.65(3.92_19.07) | <0.001** |
| Reproductive health related information in school | Yes | 254(49.2%) | 103(20%) | 0.91(0.60_1.39) | 0.93(0.55_1.57) | 0.77 |
|  | No | 116(22.5%) | 43(8.3%) | 1 | 1 | 1 |

## DISCUSSION

The overall knowledge score of this study was 71.7% CI 95%(67.6_75.4), which is lower than the studies conducted in Australia 97.4%[17], Zambia 98.7% [18], and higher than USA47.5%[19], china 15.7%[13], Taiwan7.49%[20], Spain 67%[12], Grace59.1%[21], and Turkey 23.2%[22], Nigeria 23%[23], Serbia 14.2%[24], Ghana 40%(14), and Ethiopia(AddisAbaba13% [25], and Jimma 43.8%[11]. These dissimilarities may be due to the differences in culture and information access, differences in education institutions’ structure, sampling procedures and tools, and the cut-off points to define the level of knowledge.

The age of respondents had a positive association with knowledge. Those who are >15 years old have close to fourfold more likely to have good knowledge about HPV vaccine as compared to those who are 12_14 years old, AOR =3.74, 95%CI(2.20_6.37), P-value <0.001), which is similar to a study conducted in Greek [21] and Jimma, Ethiopia[11].

As the education level of girls increases, knowledge of the HPV vaccine increases. Those who are in Grade7and 8 have close to four-fold more likely of having good knowledge about HPV vaccine as compared to those who are in Grade 5 and 6, AOR=3.98, 95%CI (2.40_6.58), P-value <0.001), a study conducted in Jimma, southwest, Ethiopia,[11] and Turkey[22] showed a similar result. The association might be due to the increased level of education that may pave the way to increased information access, and the level of understanding.

Familial educational status(those for whom, mothers educational status of secondary close to fourteen fold and more than secondary close to twenty-two fold and fathers educational status of more than secondary close to twofold have good knowledge about HPV vaccine than their counterpart, AOR=13.60, 95%CI (5.69_32.53)P-value <0.001), 22.27, 95%CI(8.23_60.30), P-value<0.001) 2.18, 95%CI(1.09_4.35)P-value 0.03) respectively, this is similar with a study conducted in Senegal [26]. It could be because of good communication between parents and their daughters and that may aware them.

Information source about HPV vaccine has a statistically significant association with knowledge of HPV vaccine, that is, those who obtained HPV vaccine information have close to nine-fold more likely of having good knowledge about HPV vaccine than those who do not obtain HPV vaccine information AOR=8.65, 95% CI(3.92_19.07), P-value <0.001) which is similar to a study conducted in USA [19].

### Strength of the study

As the study was conducted in a multidisciplinary way, the quality of the study was unquestionable. In addition to this, a pretest was conducted and relevance, consistency, and validity of the tool were checked. It was able to survey a random sample of female students, thereby increasing the generalizability of the findings to other eligible students. This study utilized a self-administered questionnaire which increases the likelihood of respondents answering openly, hence increase internal validity.

### Limitations of the study

The limitation of this study was not included girls’ parents in the study as they were decision-makers.

## CONCLUSSION

The overall knowledge level of study participants was 71.7%, which is still less than three fourth.

Factors positively associated with knowledge of HPV vaccine were age, Grade level, and familial educational status, and information source. It is better if health care workers provide extra information to increase girls’ knowledge about the HPV vaccine. It is also better if education institutions encourage girls’ education as the increased level of girls’ education is associated with the increased level of HPV vaccine knowledge.

## Data Availability

All data produced in the present work are contained in the manuscript.

## Data Availability

The data supporting the conclusions of this study were included in the text.

## ACKNOWLEDMENTS

We want to forward our respectful gratitude to the Arbaminch city education bureau for their cooperation in giving us important information.

We also want to forward our uncountable thank to directors and teachers of schools that were selected for our study as well as data collectors and students who were participated in our study by filling the questionnaires allocated to them.

## Authors’ contributions

Eshetu Yisihak made substantial contributions to the conception and design of this study. Eshetu Yisihak together with Feleke Gebremeskel made contributions to analyze and interpret the data. Desta Markos, Gebresilasea Gendisha & Samuel Abebe made contributions by guiding the collection of data and checking the consistency and completeness of the data. All authors were responsible for drafting the manuscript.

## Ethical Considerations

Ethical clearance was obtained from the University of Arba Minch, institutional ethical review board. Co-operation letter received from Gamo zone education office, and selected schools. Ascent consent from homeroom teachers of adolescents and informed consent from adolescents was obtained before the interview was commenced. The ascent was obtained from homeroom teachers of adolescents through sending ascent requesting letters from the University of Arba Minch to the schools selected for the study. Confidentiality was maintained by omitting their name and personal identification.

